# Forecasting the Number of Coronavirus (COVID-19) Cases in Ethiopia Using Exponential Smoothing Times Series Model

**DOI:** 10.1101/2020.06.29.20142489

**Authors:** Teshome Hailemeskel Abebe

## Abstract

The main objective of this study is to forecast COVID-19 case in Ethiopiausing the best-fitted model. The time series data of COVID-19 case in Ethiopia from March 14, 2020 to June 05, 2020 were used.To this end, exponential growth, single exponential smoothing method, and doubleexponential smoothing methodwere used. To evaluate the forecasting performance of the model, root mean sum of square error was used. The study showed that double exponential smoothing methods was appropriate in forecasting the future number ofCOVID-19 cases in Ethiopia as dictated by lowest value of root mean sum of square error. The forecasting model shows that the number of coronavirus cases in Ethiopia grows exponentially. The finding of the results would help the concerned stakeholders to make the right decisions based on the information given on forecasts.

## 1. Introduction

On March 11^th^2020, World Health Organization (WHO) declared the 2019 novel coronavirus as global pandemic. Coronavirus, also known as COVID-19 was first originated in Wuhan, Hubei province in China around December 2019 and spread out all over the world within few weeks (1).Following the outbreak, the World Health Organization (WHO) declares the outbreak as Public Health Emergency of International Concern on 30 January, and pandemic on 11 March 2019. In June 5, 2020, more than6,603,329million cases has been reported and resulting more than 391,732 deaths across the world.

The first case of COVID-19 pandemic was confirmedin Ethiopia on 13 March 2020. Consequently, on 16 March 2020, the government of Ethiopia announced that schools, sporting events, and public gathering were suspended for 15 days. Due to the continuation of the outbreak, on April 2020, the council of ministers declared a five-month state of emergency in response to the growing number of coronavirus cases.

Almostall-pandemic disease exhibits their own patters, which require to be defined by the level of transmission and coverage.Followingtheoutbreak, the coronavirus (COVID-19)have a fast transmission natureand grow exponentially across the globe.

subsequently, to model the exponential growing rate of the virus, different researchers (2, 3, 4)conducted their study usinga linear based time series model (Auto Regressive Moving Average (ARIMA family) models. However, such linear based time series modelscannot handle a data having anexponentialgrowingpatternandresultsfail to account the dynamics of transmission of the coronavirus. Therefore, ARIMA family models are unable to fit the data well given an exponential growth of COVID-19 transmission.

Thus, we should to find a model, which can capture a data that have an exponential growing pattern.Therefore, in this study, we use among the common exponential family models such as an Exponential Growth Model, Simple (Single) Exponential Smoothing (SES), and Double Exponential Smoothing (DES) methods.

The main motivation of the study is to identify an appropriate model for coronavirus(COVID-19) which has an exponentially increasing pattern. As a result, finding a model that capture such exponentially increasing pattern of the data is the primary motive of this study.

The rest of this paper is organized as follows: section II describes dataset and methodology. Section III presents the results and discussions and on section IV, concussionswerepresented.

## 2. Data and Methodology

### 2.1. Data

The daily data on coronavirus disease, also known as COVID-19 confirmed case in Ethiopia is collected from World Health Organization (WHO) database over the period between 14-03-2020 and 05-06-20 (5).

To develop an appropriate forecast of the data, the data set should be divided into in-sample and out-sample forecasts. Thus, the COVID-19 dataset is divided into training set (80%) on which our models are trained and testing set (20%) to test the performance of the model.

### 2.2. Model Specifications

A time series is a sequence of observations on a variable taken at discrete intervals in time. We index the time periods as 1, 2,…, T and denote the set of observations as (*Y*_l_, *Y*_2_,….,*Y*_*T*_). In time series, each element of the series is treated as a random variable with a probability distribution. Moreover, it is known that the main objective of time series analysis is to forecast the future value or patterns of the series. In this study, we try to forecast the future value of COVID-19 cases in Ethiopia based on the nature of the series.

In order to forecast the number of positive COVID-19 casesin Ethiopia, the following modelsare defined.

#### 2.2.1. Exponential Growth Trend Model

Single exponential growth model is one of the best time series model in forecasting data that have growing nature by accounting an exponentially growing (decaying) data.

Suppose a variable *Y*_*t*_ is an exponential function of time (t), then the modelis given by:

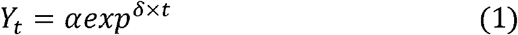

where*Y*_*t*_ is the number of COVID-19 confirmed cases, a is the initial value of *Y, δ* is a positive growth factor and *t* is the time constant required for *Y* to increase by one factor of ***δ***.

In order to estimate the exponential growth model using OLS, we should to linearize it using a log transformation.

Therefore, the linearize trend model is given by:

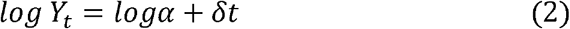

The exponential growth model can be used to estimate the parameters as well as to forecast the future cases of COVID-19. For forecasting part, we can compare it with exponential smoothing (single and double) methods since they are also appropriate to forecast data that have an exponential pattern. Thus, we can compare the forecasting performance of exponential growth model and exponential smoothing methods. Therefore, the single and double exponential smoothing techniques are discussed below.

#### 2.2.2. Simple(Single) Exponential Smoothing (SES)

Exponential smoothing was introduced in the late 1950s. Exponential Smoothing is a method of smoothing time series data based on the exponential window function. The exponential functions are used to assign exponentially decreasing weights over time.

It is method of data analysis obtained by using some optimal weight generated according to the data estimations with a given specific weight.Forecastsproduced using exponential smoothing methods weighted averages of past observations. These methods give decreasing weights to past observations and thus the more recent the observation the higher the associated weight. This framework enables reliable estimates to be produced quickly in most applications. Single Exponential Smoothing method is used when the time series data has no trend and no seasonality.

The smoothing function for any time period t as defined by (6)is given by:

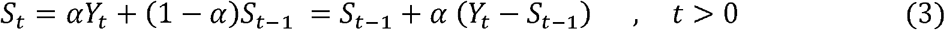

where*S*_t_ denotes the current smoothed series obtained by applying simple exponential smoothing series Y.*Y*_*t*_is the current observed value of the time series in period *t, α* is the smoothing constant or factor ranging from 0 to 1, 1− *α* estimates the moving average parameter and *s*_*t*−l_ is the smoothed value at time *t* − 1. In exponential smoothing technique, *S*_*t*_ = *Y*_*t*_.

A value of *α* close to one have less of a smoothing effect and gives a greater weight to recent changes in the data, while values of *α*closer to zero have a greater smoothing effect and are less responsive to recent changes.

The *h* − *step* − *head* prediction is:

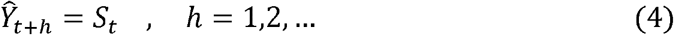

The general form of single exponential smoothing forecast function is:

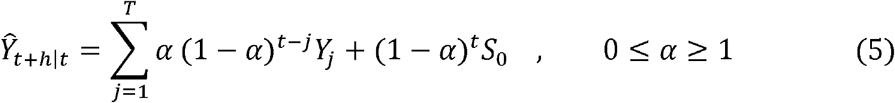

where*h* is the number of periods in the forecast lead-time and *Ŷ*_*t*+*h*_ is the forecast for *h* periods ahead from origin *t*.

#### 2.2.3. Double Exponential Smoothing

Single exponential smoothing cannot smooth well when there is trend in the data. Thus,we should to use a double exponential smoothing (second order exponential smoothing) method when the time series data has a trend but not seasonality component.

The smoothing function for any time period t isgiven by:

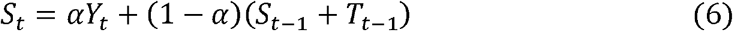

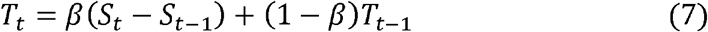

wherethe first equation refers to the level, while the second equation is the trend. *S*_*t*_ denotes an estimate of the level of the series at time *t, T*_t_ is the smoothed additive trend at the end of period *t,α* is the data smoothing parameter for the level of the series and*β* is the smoothing parameter for the trend range between (0,1).

The *h* − *Step* − *head* prediction is:

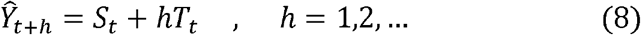

### 2.3. Model Diagnostic to choose the best forecasting model

According to (7) forecasting is an important application of time series analysis. However, theperformance of forecasting a given model should be evaluated by using different evaluation criteria. Among the common evaluation criteria is therootmeansum of square errors (RMSSE).The models, which have the smallest root mean sum of square error,will be used as the best forecasting model.

The root mean sum of square error is given by:

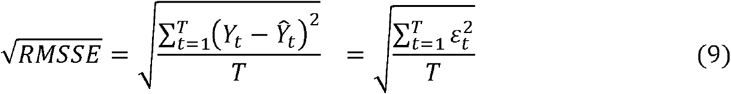

where Ŷ_*t*_ is the expected (forecasted) value for period t, *Y*_*t*_ is the actual (observed) value for the period t, and T is the number of periods.

## 3. Results and Discussions

To analysis the dataset, the researcher uses Stata 13 for exponential growth analysis and Microsoft excel 10 for single and double exponential smoothing method analysis due to the simplicity of the software for the respected models. Therefore, if you look some difference on the Figure format, that is a matter ofsoftware package difference.

### 3.1. Time Plot of the Data

From Figure 1, the horizontal line shows the number of days and the vertical axis is the total number of COVID-19 cases in Ethiopia. From the figure, we observe that the series grow exponentially.

**Figure 1:**
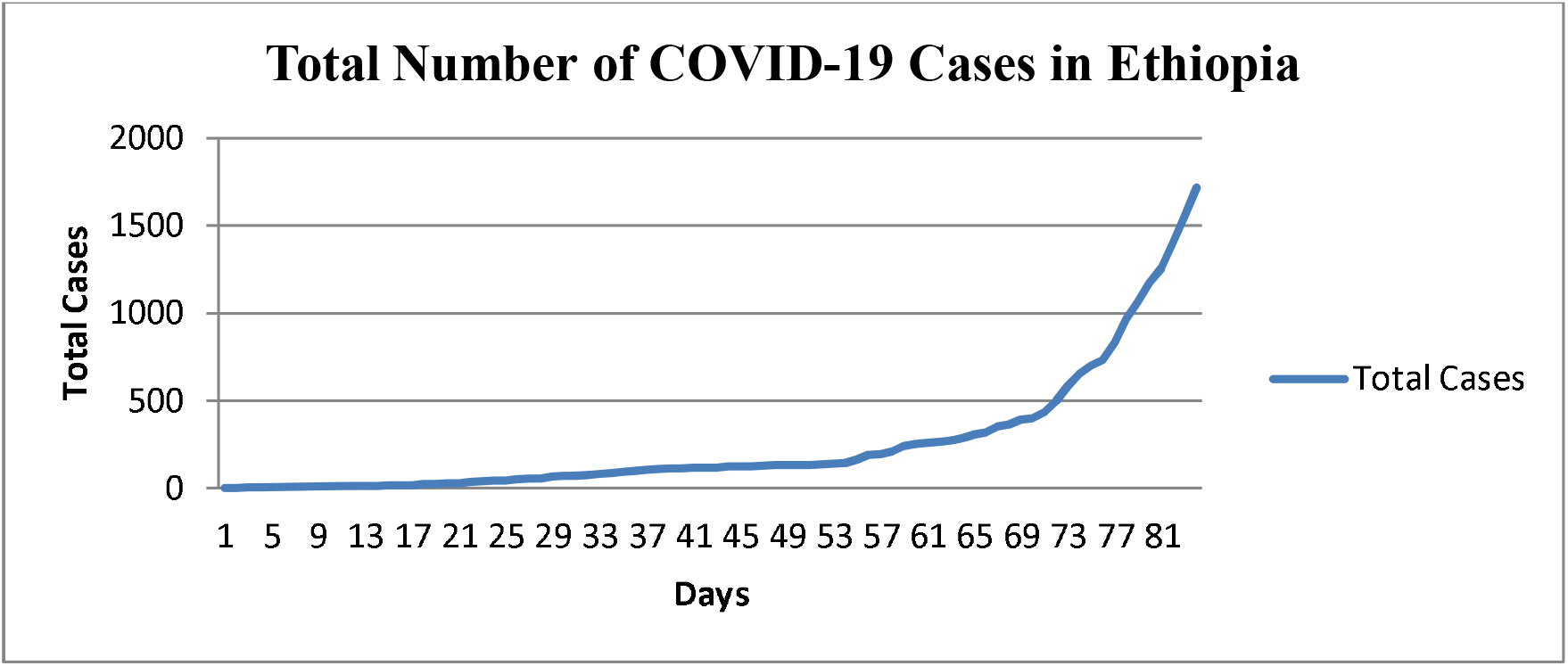
Time series line on the original series Source: Author’s Computations

### 3.2. Estimation model Results

In order to identify an appropriate model for forecasting the number of COVID-19 cases in Ethiopia, an exponential growth, single and double exponential smoothing methods are used as a candidate model.After fitting the three models, the following results are obtained.

As shown in Table 1, wehave established the following time series techniques for analyzing and forecasting the number of confirmed cases in Ethiopia. When we compare the three competitive candidate models, we find that adouble exponential smoothing technique is better than the other models as indicated by the highest coefficient of determination (R2) and F-statistic test.Therefore, according to Table 1, we find that double exponential smoothing technique is the best of the other two time series models in terms ofgoodness of fit criteria.

**Table 1:** Summary of fitted time series models for total confirmed COVID-19 cases in Ethiopia

| Methods | $R^2$ | F-test | P-value |
| --- | --- | --- | --- |
| Exponential Growth Model | 0.8871 | 99.82 | 0.000 |
| Single exponential smoothing Model | 0.9296 | 174.95 | 0.000 |
| Double exponential smoothing Model | 0.9577 | 181.5 | 0.000 |

Source: Author's Computations

### 3.3. In-Sample Prediction Using Graphically

From the Figure, we observe that, the difference between the actual value and the forecast value in double exponential smoothing method is relatively smaller than that of exponential growth and simple exponential smoothing methods. Thus, the double exponential smoothing method in a better way to predictthe number of COVID-19 cases in Ethiopia on the coming three weeks.

### 3.4. Model Diagnostic

Beside the graphical evaluation of the performance of the fitted model for a given series, we can evaluate it using formal statistical methods of evaluation. The performance of the model is depending on how the actual value is close to the predicted value. In this case, the root mean sum of square error (RMSSE) is used to evaluate the performance of the three models (exponential growth, single exponential and double exponential models). Among the three models, the double exponential smoothing model was the best-fittedmodelfor analysis and forecast the number of COVID-19 cases in Ethiopia as given by small values of RMSSE. Therefore, this model is best-fitted model and selected as a best model for forecasting the future cases of the COVID_19 cases. The diagnostic measures for the selection of best forecasting model is given by Table 2.

**Table 2:** Diagnostic measures for selection of best-fitted model

| Forecasting Models | RMSSE |
| --- | --- |
| Exponential Growth model | 0.21839 |
| Simple Exponential Smoothing Model | 0.41142 |
| Double Exponential Smoothing Model | 0.03978 |

Source: Author's Computations

### 3.5. Forecasting

Forecasting is making prediction of some future value of events using past and present data.However, as suggested by Neils Bohr, making good prediction is not always easy (8). Moreover, most statistical time series methods are valid for short term forecasting (such as days, weeks, months) and medium term forecasts (one to two years) than long term forecasts (more than two years) since most historical data usually exhibit inertia and do not change dramatically very quickly.

From the model diagnostic result on Table 2, the best-fitted model based on minimum root mean sum of square error (RMSSE) is, a double exponential smoothing method. Therefore, to forecast the future COVID-19 cases, a double exponential smoothing method given in Figure 5 as below.

**Figure 2:**
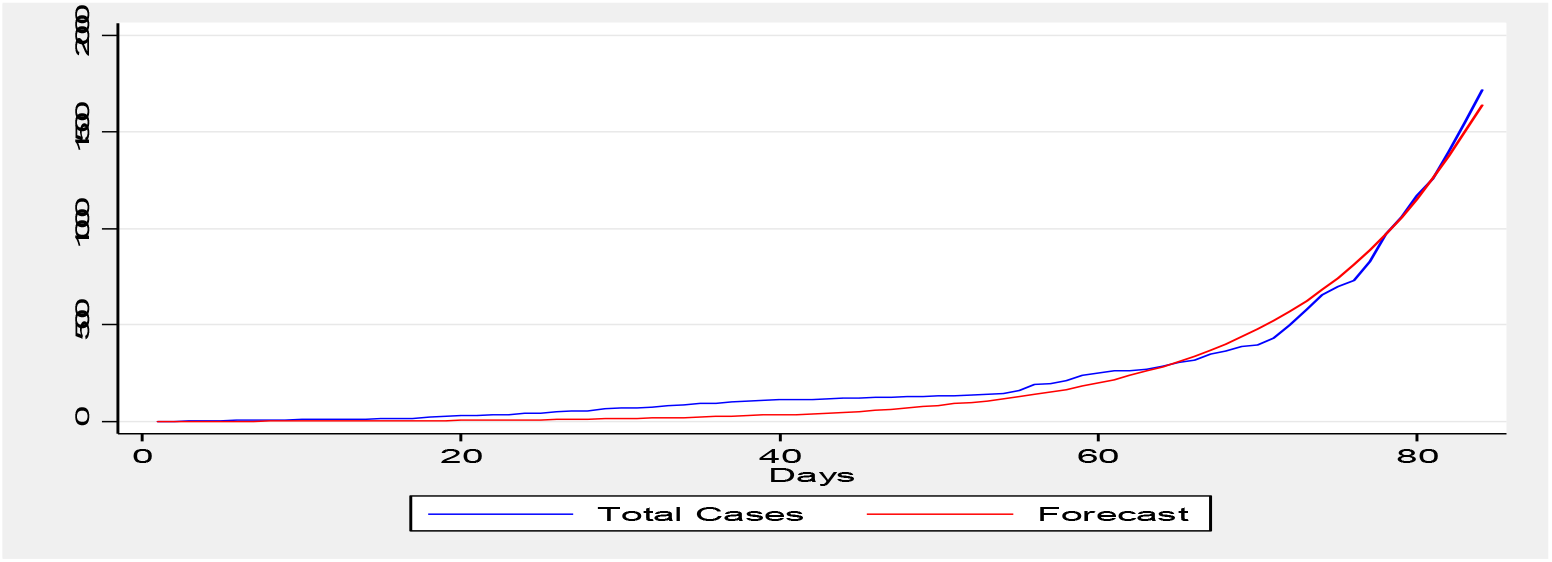
In-Sample Prediction Using Exponential Growth Source: Author’s Computations

**Figure 3:**
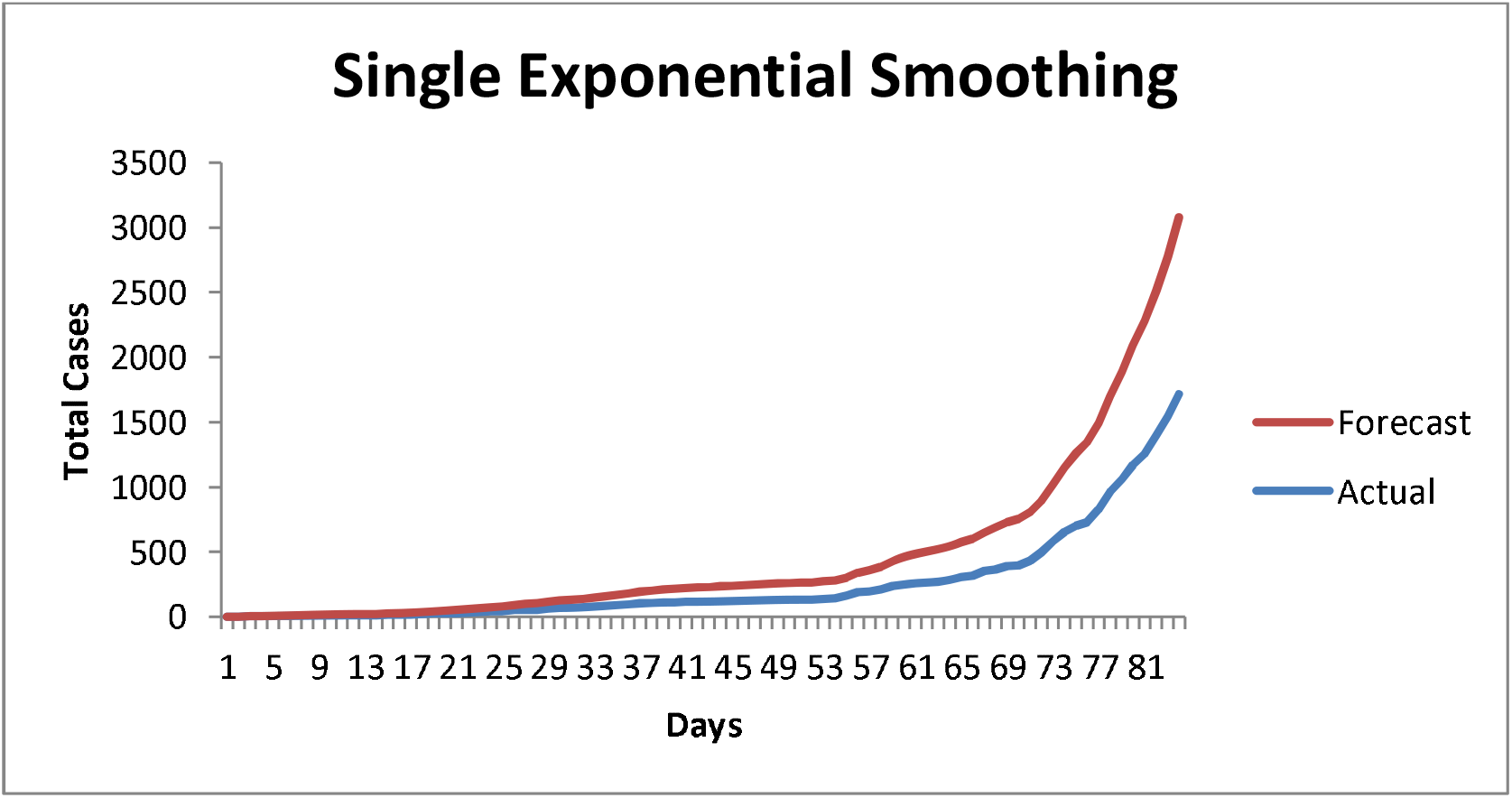
In-Sample Prediction Using Simple Exponential Smoothing Source: Author’s Computations

**Figure 4:**
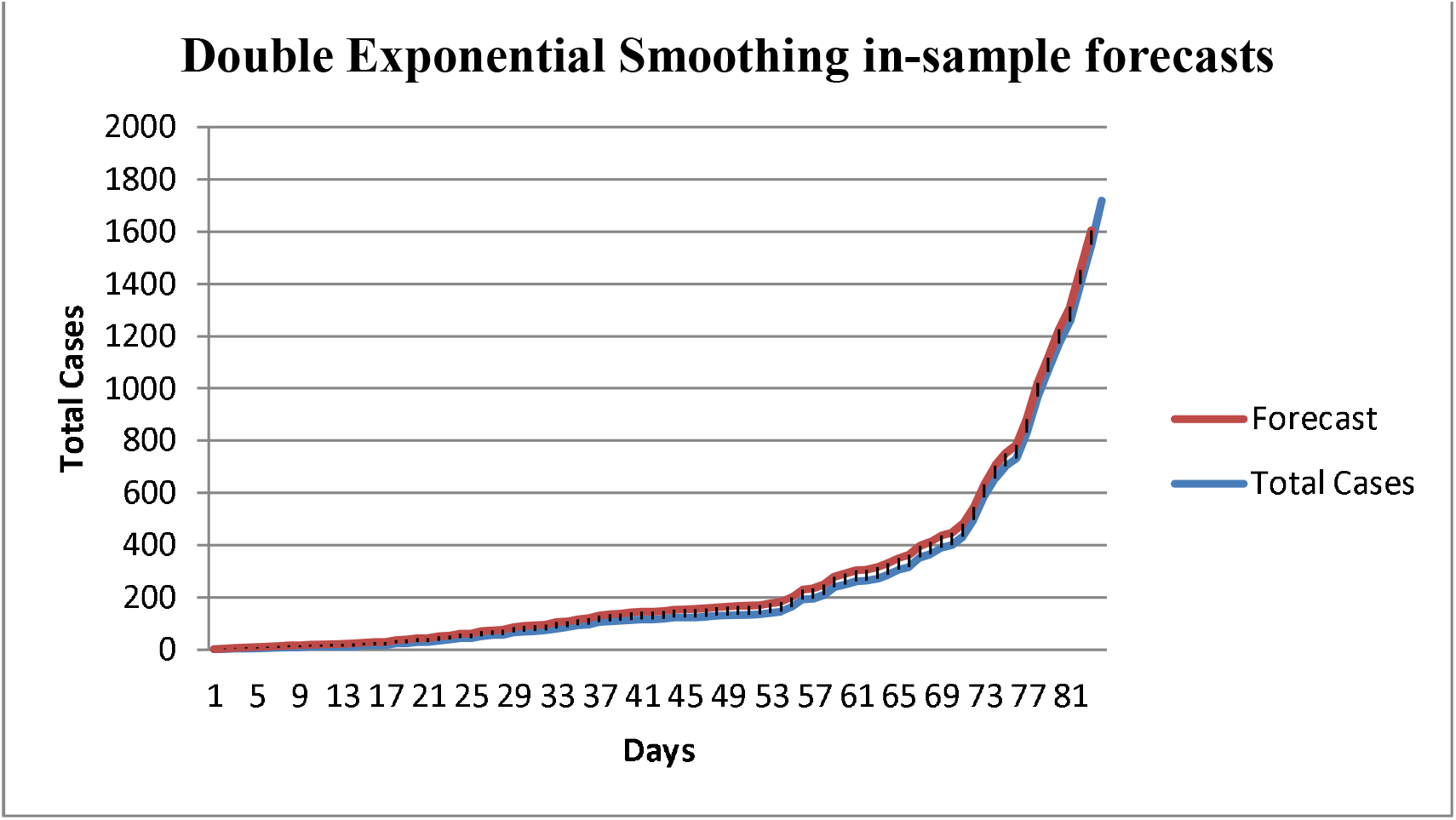
In-Sample Prediction Using Double Exponential Smoothing Source: Author’s Computations

**Figure 5:**
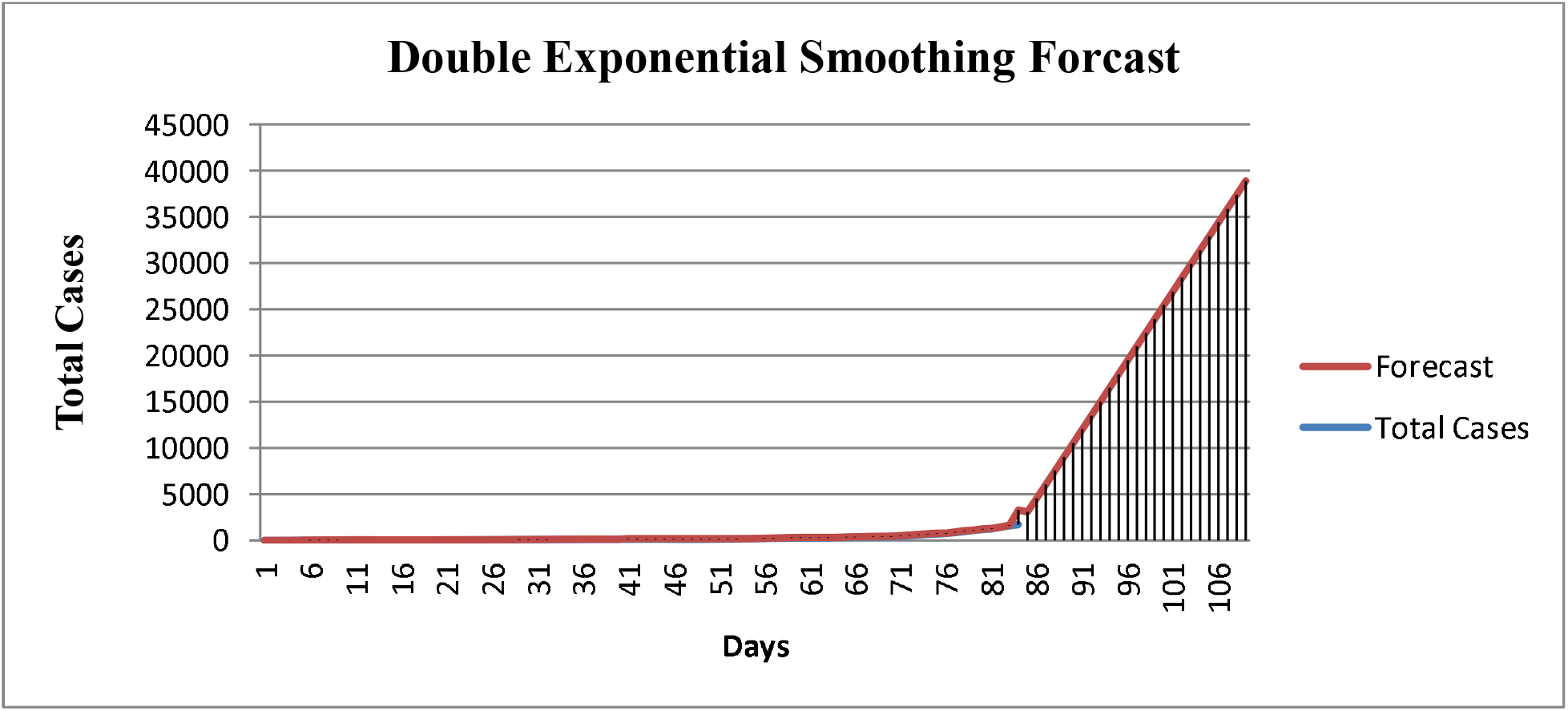
Forecasts Using Double Exponential Smoothing Methods Source: Author’s Computations

## 4. Conclusions

In this study, an exponential growth, simple and double exponential smoothing methods were fitted on COVID-19 data of Ethiopia from the period 14-03-2020 to 05-06-2020.

The forecasting performance of the model is evaluated using Root Mean Sum of Square Error (RMSSE). The result shows that double exponential smoothing method is better than other two modelsinforecast future cases of COVID-19 in Ethiopia.From the result of forecast, the number of people who will be affected by COVID-19 in Ethiopia increasesin an exponential manner in the next three weeks.

Thus, the forecast helps the Ethiopian government, policy makers, and the society at alltotake preventive measures before the transmission become out of control especially rural areas since until now, most of the cases were observed in urban areas.

## Data Availability

I will send in case of need

## Abbreviations and Acronyms

ARIMA: Autoregressive Integrated Moving Average
DES: Double exponential smoothing
OLS: Ordinary least Square
RMSSE: Root Mean Sum of Square error
SES: Single exponential smoothing
WHO: World Health Organization

## Author contributions

Since Mr. Teshome is the only author of this paper, his contribution is not only on formulating the idea of the research, but also in collecting, analyzing, and interpretation of the data. Thus, the author read and approved the final manuscript by himself.

## Conflicts of interests

The author declares no conflict of interest.

## Funding

This research did not receive any fund.

## Notes

### Competing Interest Statement

The authors have declared no competing interest.

### Clinical Trial

because i am not health professional

